# Real-World Comparison of Clopidogrel with Ticagrelor and Prasugrel In Patients with Chronic Coronary Disease undergoing Atherectomy

**DOI:** 10.1101/2023.10.24.23297498

**Authors:** Mahin R. Khan, Anoop N. Koshy, Richard Tanner, Serdar Farhan, Ali Farooq, Samantha Sartori, Yihan Feng, Alessandro Spirito, Ayush Arora, Vishal Dhulipala, Manish Vinayak, Vishal Kapur, Javed Suleman, Raman Sharma, Roxana Mehran, Annapoorna Kini, Samin K. Sharma

## Abstract

**Background:** In patients with chronic coronary disease (CCD), it is unclear whether the use of potent P2Y_12_ inhibitors (ticagrelor or prasugrel) offers advantages to clopidogrel when prescribed in conjunction with aspirin in patients undergoing percutaneous coronary intervention (PCI) with atherectomy.

**Methods:** Consecutive patients undergoing PCI with atherectomy for CCD at a tertiary care center between January 2011 to December 2020 were included. Patients discharged on ticagrelor or prasugrel were compared to patients on clopidogrel. The primary outcome was a composite of death or myocardial infarction (MI), secondary outcomes included individual components of the primary outcome, stroke, major bleeding, and target vessel revascularization at 1 year. Adjusted analyses were performed using propensity score stratification.

**Results:** Overall, 3,612 patients undergoing atherectomy were included in the analysis (clopidogrel [70.4%, n= 2,543], ticagrelor/prasugrel [29.5%, n=1,069]). Clopidogrel was prescribed more often in older patients with multimorbid risk factors, whereas ticagrelor/prasugrel was used more in patients with greater anatomical and procedural complexity. There was an increase in the use of potent antiplatelet agents over time (p<0.001). At 1-year follow-up, the primary outcome was observed in 5.2% and 4.0% of those taking clopidogrel and ticagrelor/prasugrel, respectively (adjusted hazard ratio (AHR) 0.87, 95% CI 0.58 – 1.3, p = 0.50). There were no significant differences in the rate of bleeding (5.5% vs 3.7%, AHR 0.98, 95% CI 0.66 - 1.46, p = 0.92) or other secondary outcomes between the two groups.

**Conclusion:** The use of clopidogrel was associated with comparable ischemic and bleeding outcomes compared to ticagrelor/prasugrel in patients with CCD undergoing PCI with atherectomy.

## INTRODUCTION

Dual antiplatelet therapy (DAPT) is recommended for patients after percutaneous coronary intervention (PCI), with the choice of antiplatelets being guided by the patient’s clinical presentation rather than patient factors or the complexity of the coronary intervention. Current societal guidelines reserve potent P2Y12 platelet receptor inhibitors (ticagrelor, prasugrel) for acute coronary syndrome (ACS) cases due to a paucity of evidence for their use in chronic coronary disease (CCD).^1^

Patients with CCD undergoing PCI with adjunctive atherectomy represent a subset of patients with more complex coronary disease who may derive benefit from more potent platelet inhibition. Atherectomy is associated with an increased risk of microvascular injury, endothelial dysfunction, and platelet activation which may contribute to the increased risk of thrombosis, myocardial infarction and death seen in this patient cohort.^2^ However, it is not known if these complications can be mitigated by the use of potent inhibitors rather than clopidogrel.

Despite a paucity of evidence supporting the use of potent P2Y12 inhibitors in CCD, real-world data has demonstrated that their off-label use is continuing to increase. ^3^ Given the potential for a higher risk of bleeding with these agents and the higher cost to patients there is an urgent clinical need to further scrutinize their use CCD. ^4,5^ It is plausible that higher complexity CCD cases, such as those treated with atherectomy may benefit from the use of ticagrelor and prasugrel. Therefore, we sought to compare the clinical outcomes of CCD patients treated with PCI with adjunctive atherectomy prescribed DAPT comprising of aspirin plus clopidogrel or aspirin plus ticagrelor or prasugrel.

## METHODS

### Study population and design

Consecutive patients undergoing PCI at Mount Sinai Hospital, New York from January 2011 to December 2020 and enrolled in the institutional database were considered for inclusion. All patients undergoing PCI for CCD discharged on DAPT were subsequently included for analysis. For the purpose of this analysis, patients were divided into two groups based on whether they received a potent P2Y_12_ (ticagrelor or prasugrel) or clopidogrel in addition to aspirin. Exclusion criteria included patients presenting with non-ST segment myocardial infarction (NSTEMI) or ST-segment myocardial infarction (STEMI), cardiogenic shock, and those on oral anticoagulation. In keeping with previous studies from our institution, event-free patients lost to follow-up within 30 days after the index procedure were excluded.

Baseline and procedural data were prospectively collected. Our standard follow-up protocol encompasses mandatory contact with patients at 30-days and 1 year after PCI (in-person or telephone) by dedicated research staff. Additional supporting data was gleaned from hospital medical records, by contacting primary care physicians, and through external death records (on-line obituary and Social Security Death Index). All patients provided written informed consent before PCI was performed and the study was approved by the Institutional Board Review at Mount Sinai Hospital, New York.

A decision to perform atherectomy and the type of atherectomy device used was at the discretion of the operator and guided per lesion morphology. At this institution, rotational atherectomy (RA) and orbital atherectomy (OA) are the devices of choice for moderate to severe calcification modification and Excimer laser coronary atherectomy (ELCA) is reversed for balloon uncrossable lesions.

This study was approved by the Institutional Board Review at Mount Sinai Hospital, New York. The data that support the findings of this study are available from the corresponding author upon reasonable request.

### Antiplatelet regimen

All patients at this institution receive 325 mg aspirin more than 90 minutes prior to angiography, followed by a maintenance dose of 81 mg daily. Once a decision is made to proceed with PCI, patients are administered a P2Y_12_ inhibitor (clopidogrel/ ticagrelor/ prasugrel) in the catheterization laboratory if not already taking one. Loading doses for each P2Y_12_ inhibitor are standard; ticagrelor 180mg, clopidogrel 600mg, prasugrel 60mg. As per institutional protocol a maintenance dose of prasugrel 10mg was administered when body weight was >100kg or if 3 stents were deployed in the same vessel, all other patients on prasugrel received 5mg once daily after loading. Maintenance doses of ticagrelor and clopidogrel were 90mg twice daily and 75mg once daily respectively.

Our standard practice is to prescribe a P2Y_12_ inhibitor in addition to aspirin for 12 months after PCI for CCD. Aspirin monotherapy is continued after the 12 months of DAPT. Routine escalation or de-escalation of antiplatelet therapy was not performed.

### Atherectomy protocol

RA was performed using the Boston Scientific Rotablator System as previously described. ^6^ Briefly, individual RA burring runs were strictly limited to 15 to 20 seconds with burr speeds of 140,000 to 150,000 rpm and targeted overall burr-to-artery ratio of 0.5. Rotaflush with normal saline containing a cocktail of nitroglycerine (5 mg/L), verapamil (5 mg/L), and heparin (5000 U/L) was continuously infused through the Teflon rotablator sheath throughout the procedure. The Diamondback 360° Coronary Orbital Atherectomy System 1.25 mm Classic Crown (Cardiovascular Systems, Inc. [CSI], St. Paul, MN) was used in all cases. The OA system was advanced over the ViperWire (CSI) with a rotational speed of 80,000 rpm (low speed) or 120,000 rpm (high speed) depending on vessel size. In our institutional practice, OA is started at low speed and high speed is only used when additional debulking is required. ViperSlide lubricating solution (CSI) is infused into the device during the procedure. The CVX-300 Excimer Laser System (Spectranetics Inc., Colorado Springs, CO), in conjunction with the 0. 9-mm X80 catheter (Spectranetics Inc.), was used. The fluence, defined as the amount of energy (mJ) at the catheter tip per area unit (mm2), usually has a range between 30 and 80 mJ/mm2 and the repetition rate (frequency) range is between 25 and 80 Hz (pulses/s) which was chosen at operator discretion. Reaching the maximum energy and rate is standard practice if the resistance to pass the lesion persisted and flushing with continuous saline was performed while 10-s pulses were delivered ^7^.

### Study definitions and endpoints

The primary endpoint of the study was the composite of major adverse cardiovascular events (MACE), defined as all-cause death or myocardial infarction (MI) at 1 year following PCI. Secondary endpoints included the individual components of the primary endpoint, target lesion revascularization, stent thrombosis, major bleeding events and cerebrovascular accident (CVA). MI was defined according to the third universal definition of MI ^8^. TLR was defined as repeat revascularization within the 5 mm margin proximal and distal to the stent. Bleeding events were defined as per the National Cardiovascular Data Registry CathPCI Registry (version 4.4), including any bleeding occurring during hospitalization associated with a decrease in hemoglobin >3 g/dL, blood transfusion, procedural intervention or surgery at the bleeding site, or bleeding requiring hospitalization or a blood transfusion. Complex PCI was defined based on predetermined criteria, including total stent length ≥60 mm, total number of lesions ≥3, total number of target vessels ≥3, bifurcation lesion with ≥2 stents, or chronic total occlusion (CTO) ^9^.

### Statistical analysis

Descriptive statistics were used to summarize continuous variables as mean ± standard deviation or median (interquartile range), and categorical variables were presented as frequencies and percentages. To compare baseline characteristics between the ticagrelor or prasugrel group and the clopidogrel group, the Student’s t-test, Mann-Whitney U test, or chi-square test was performed as appropriate. Trends in the use of antiplatelets over the study duration were estimated using the Cochran-Armitage test. Predictors of antiplatelet prescription were analysed by stepwise logistic regression with backward selection (p<0.10 for inclusion in the multivariate model and p>0.20 for exclusion) with risk estimates presented as odds ratios (OR) with 95% confidence intervals (CIs). Survival curves were generated using the Kaplan-Meier method, and differences between groups were assessed using the log-rank test.

To address the differences in baseline characteristics between patients who were prescribed clopidogrel vs ticagrelor or prasugrel, we conducted a propensity score stratification analysis. The propensity scores were calculated using a multivariable logistic regression model, where the outcome of interest was the choice of treatment between clopidogrel and prasugrel or ticagrelor. The propensity model was developed iteratively following the approach described by Rosenbaum et al.^10^ Propensity score stratification was then applied to examine the outcomes using cause-specific Cox proportional hazards regression models. This approach allowed us to consider the time-to-event nature of the data and stratify the analysis based on the propensity to receive either P2Y12 inhibitor. For the Cox regression and propensity score stratification, we adjusted for several variables to account for confounding factors. These variables included age, sex, body mass index (BMI), race, smoking status, hypertension, hyperlipidemia, cerebrovascular disease, prior myocardial infarction, prior coronary artery bypass grafts (CABG), left ventricular ejection fraction, calcification, chronic total occlusion (CTO), and complex PCI. To ensure unbiased selection, the process of generating mutually exclusive strata based on propensity scores for the entire cohort was blinded to any outcome data. The distribution of propensity scores for the entire cohort and each treatment group. The number of strata and the cut points for each stratum were determined based on predefined criteria and achieving adequate balance in baseline covariates. All reported p-values were two-tailed, with p < 0.05 considered statistically significant. Statistical analysis was performed using (Stata 16 MP, TX).

## RESULTS

A total of 3,612 patients undergoing PCI with atherectomy for CCD were included in the analysis. In the overall cohort, 70.4% of patients were administered clopidogrel (n=2,543) and 29.6% (n=1,069) received ticagrelor or prasugrel in addition to aspirin. Within the potent P2Y_12_ group 56.3% (n=602) patients received ticagrelor and 43.7% (n=467) received prasugrel.

Patients in the potent P2Y_12_ cohort had a higher percentage of patients with diabetes mellitus (52.0% vs 46.6%, p=0.003), prior myocardial infarction (31.9% vs 24.0%, p<0.001) and prior PCI (65.9% vs 50.5%, p =0.003). In addition, potent P2Y12 recipients were younger (65.3±9.3 vs, 70.9±10.5, p<0.001), less likely to be female (21.8% vs. 28.1%, p<0.001), and less likely to have anemia (41.8% vs. 46.8%, p=0.007) (**Table 1**).

**Table 1.**
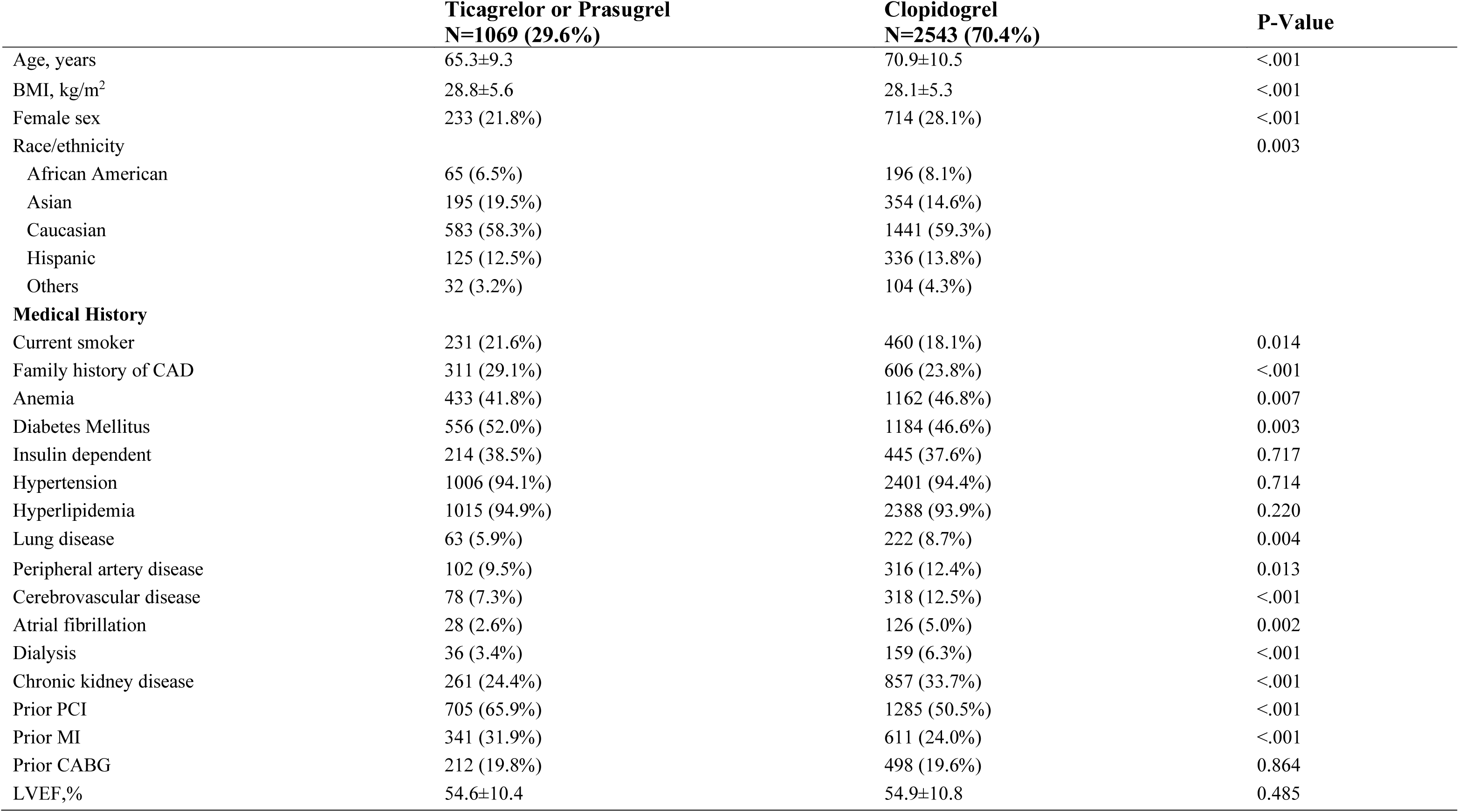
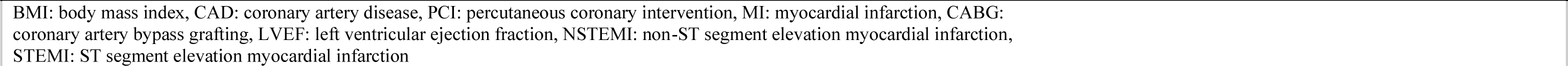
Baseline clinical characteristics.

|  | **Ticagrelor or Prasugrel**<br>**N=1069 (29.6%)** | **Clopidogrel**<br>**N=2543 (70.4%)** | **P-Value** |
| --- | --- | --- | --- |
| Age, years | 65.3±9.3 | 70.9±10.5 | <.001 |
| BMI, kg/m<sup>2</sup> | 28.8±5.6 | 28.1±5.3 | <.001 |
| Female sex | 233 (21.8%) | 714 (28.1%) | <.001 |
| Race/ethnicity |  |  | 0.003 |
| African American | 65 (6.5%) | 196 (8.1%) |  |
| Asian | 195 (19.5%) | 354 (14.6%) |  |
| Caucasian | 583 (58.3%) | 1441 (59.3%) |  |
| Hispanic | 125 (12.5%) | 336 (13.8%) |  |
| Others | 32 (3.2%) | 104 (4.3%) |  |
| **Medical History** |  |  |  |
| Current smoker | 231 (21.6%) | 460 (18.1%) | 0.014 |
| Family history of CAD | 311 (29.1%) | 606 (23.8%) | <.001 |
| Anemia | 433 (41.8%) | 1162 (46.8%) | 0.007 |
| Diabetes Mellitus | 556 (52.0%) | 1184 (46.6%) | 0.003 |
| Insulin dependent | 214 (38.5%) | 445 (37.6%) | 0.717 |
| Hypertension | 1006 (94.1%) | 2401 (94.4%) | 0.714 |
| Hyperlipidemia | 1015 (94.9%) | 2388 (93.9%) | 0.220 |
| Lung disease | 63 (5.9%) | 222 (8.7%) | 0.004 |
| Peripheral artery disease | 102 (9.5%) | 316 (12.4%) | 0.013 |
| Cerebrovascular disease | 78 (7.3%) | 318 (12.5%) | <.001 |
| Atrial fibrillation | 28 (2.6%) | 126 (5.0%) | 0.002 |
| Dialysis | 36 (3.4%) | 159 (6.3%) | <.001 |
| Chronic kidney disease | 261 (24.4%) | 857 (33.7%) | <.001 |
| Prior PCI | 705 (65.9%) | 1285 (50.5%) | <.001 |
| Prior MI | 341 (31.9%) | 611 (24.0%) | <.001 |
| Prior CABG | 212 (19.8%) | 498 (19.6%) | 0.864 |
| LVEF,% | 54.6±10.4 | 54.9±10.8 | 0.485 |

BMI: body mass index, CAD: coronary artery disease, PCI: percutaneous coronary intervention, MI: myocardial infarction, CABG: coronary artery bypass grafting, LVEF: left ventricular ejection fraction, NSTEMI: non-ST segment elevation myocardial infarction, STEMI: ST segment elevation myocardial infarction

Procedurally, the potent P2Y_12_ group had a comparatively lower proportion of patients treated via femoral access (80.2% vs 84.8%, p<0.001), had a higher percentage of left main involvement (14.2% vs 10.5%, p=0.001) and cases were more likely to involve complex PCI (64.5% vs 50.0%, p<0.001). RA was the modality of choice in >80% of cases in both groups. A higher proportion of patients were treated with RA in the clopidogrel group (85.1% vs 81.4%, p=0.005) and OA in the potent P2Y_12_ group (16.9% vs 14.0%, p=0.024). ECLA was used in less than 3% of cases for both groups, (**Table 2**).

**Table 2.**
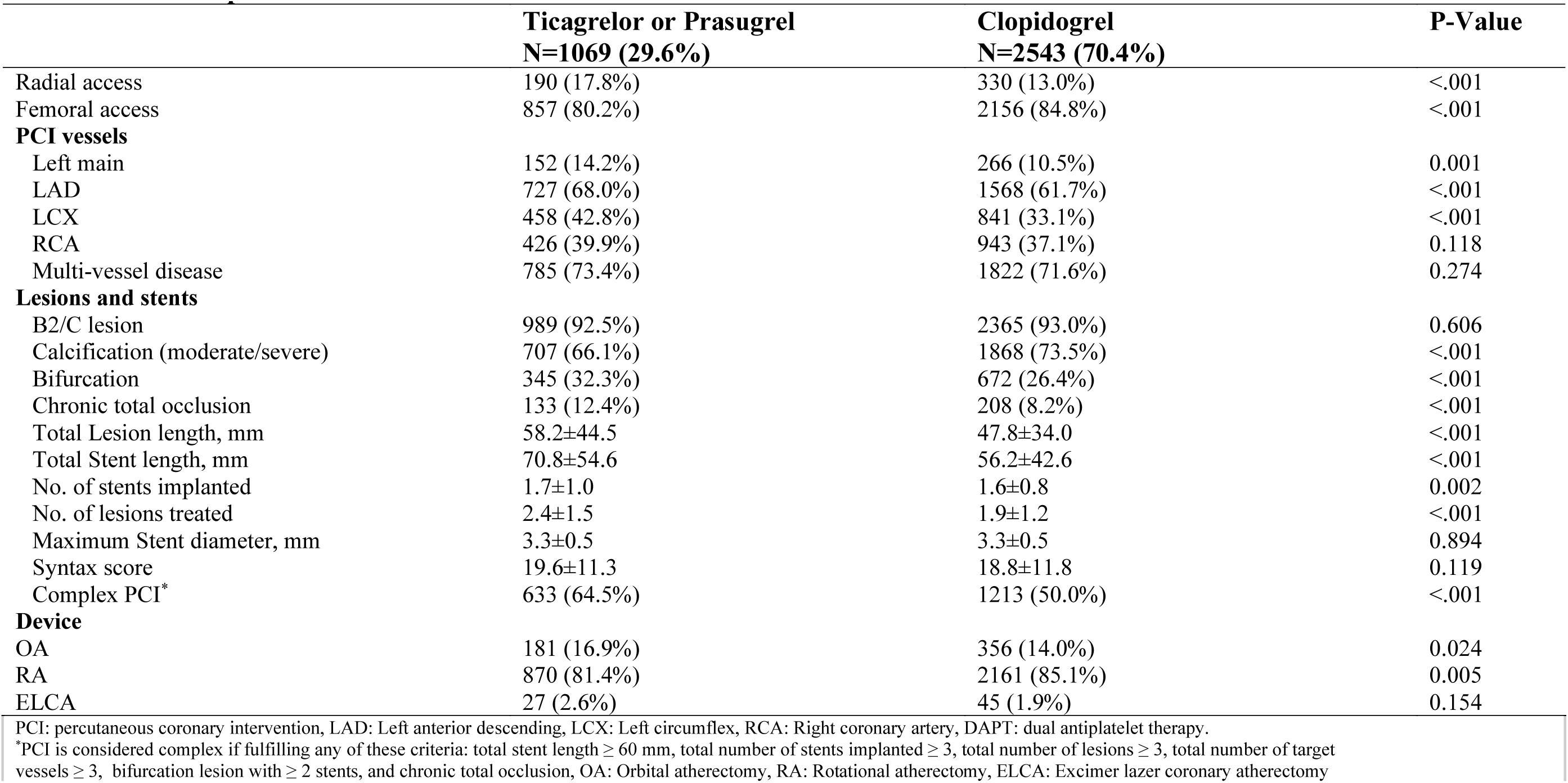
Baseline procedural characteristics.

Over the study period, the proportion of patients treated with potent P2Y_12_ inhibitors significantly increased (**Figure 1**). **Table 3** summarizes the multivariate predictors affecting P2Y_12_ therapy choice. Independent predictors of potent P2Y_12_ inhibitors included prior MI (OR 0.73, 95%CI 0.62-0.88, p=0.001) and complex PCI (OR 0.08, 95%CI 0.05-0.70, p<0.001). Conversely, clopidogrel was more commonly used with increasing age (OR 1.66, 95%CI 1.05-1.07, p <0.001) and black race (OR 1.50, 95%CI 1.08-2.09, p=0.015).

**Figure 1.**
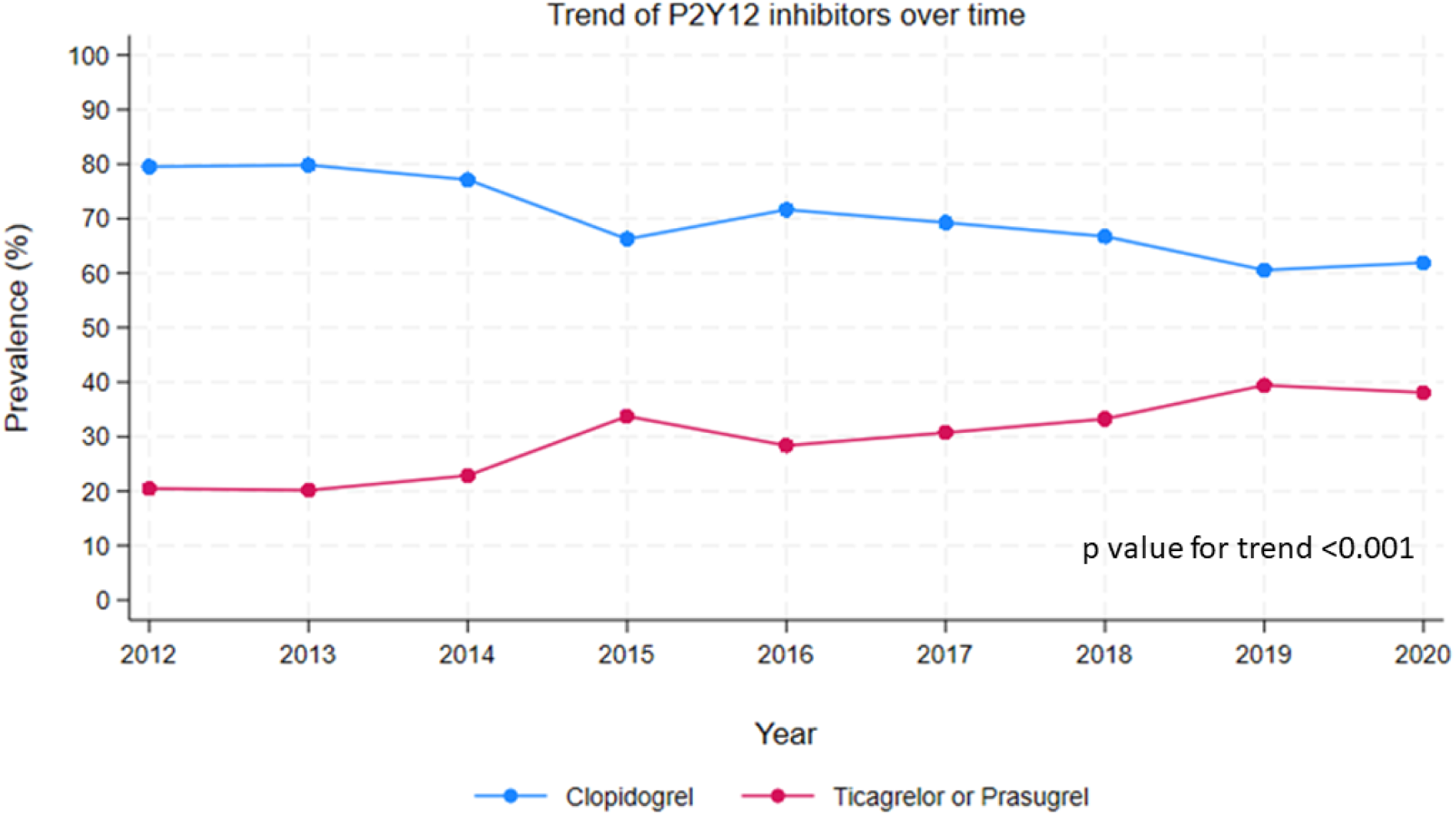
Trends in use of antiplatelets over time.

**Table 3.** Predictors for the use of Clopidogrel over ticagrelor/prasugrel.

| Predictors | HR | OR 95% CI | p-value |
| --- | --- | --- | --- |
| Age | 1.06 | 1.06 (1.05-1.07) | <0.001 |
| Race |  |  |  |
| Black | 1.50 | 1.08-2.09 | 0.015 |
| Asian | 1.01 | 0.81-1.26 | 0.909 |
| Hispanic | 1.23 | 0.97-1.59 | 0.091 |
| White | ref | - | - |
| Complex PCI | 0.08 | 0.05-0.70 | <0.001 |
| Prior MI | 0.73 | 0.62-0.88 | 0.001 |

OR: odds ratio, MI: myocardial infarction, PCI: percutaneous coronary intervention

### Clinical outcomes

Clinical outcomes at 1 year are reported in **Table 4 and Figure 3**. The distribution of propensity scores for the entire cohort and treatment group were visually examined demonstrating good overlap between groups (**Figure 2**). The primary endpoint of death or MI at 1-year follow up occurred in 38 (4%) and 112 (5.2%) of patients discharged with ticagrelor or prasugrel and clopidogrel, respectively (adjusted hazard ratio 0.87, 95%CI 0.58-1.32, p=0.518. Patients in the potent P2Y_12_ group had a lower crude risk of all-cause death (1.4% vs 2.7%, HR 0.5, 95% CI 0.27-0.91, p=0.023) on unadjusted analysis but the difference was no longer significant after propensity matching (AHR 0.73, 95% CI 0.37 - 1.42, p=0.347).

**Table 4.**
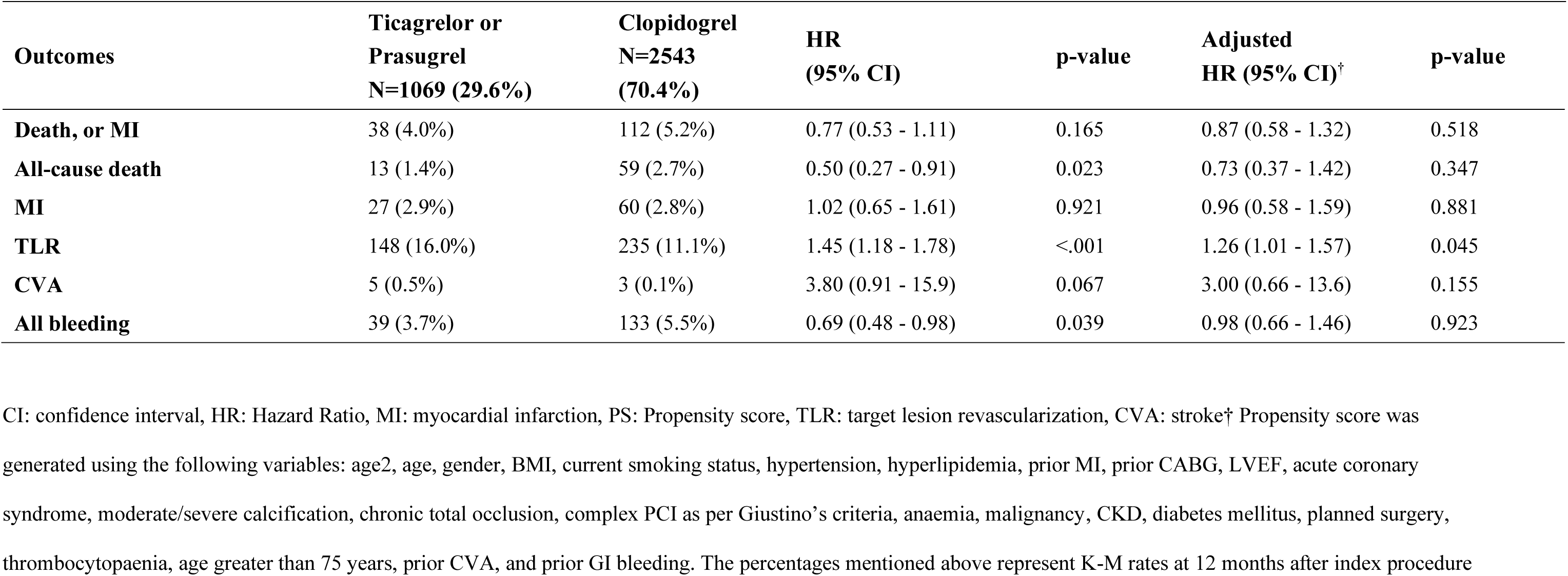
PS stratification adjusted associations between medications at discharge and adverse events.

**Figure 2.**
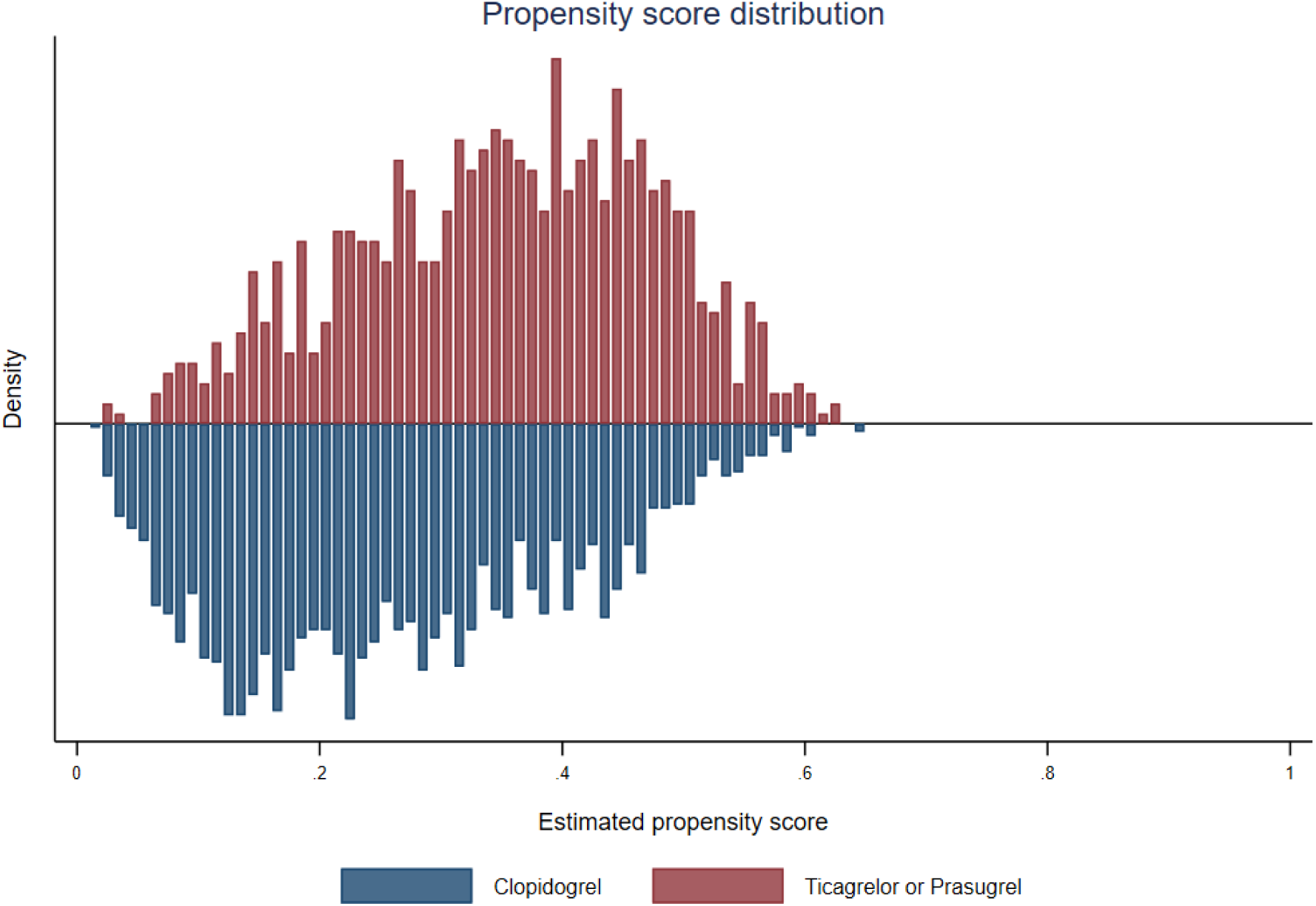
Distribution of propensity scores Propensity score distribution for patients taking clopidogrel or ticagrelor or prasugrel demonstrating good overlap between groups.

**Figure 3.**
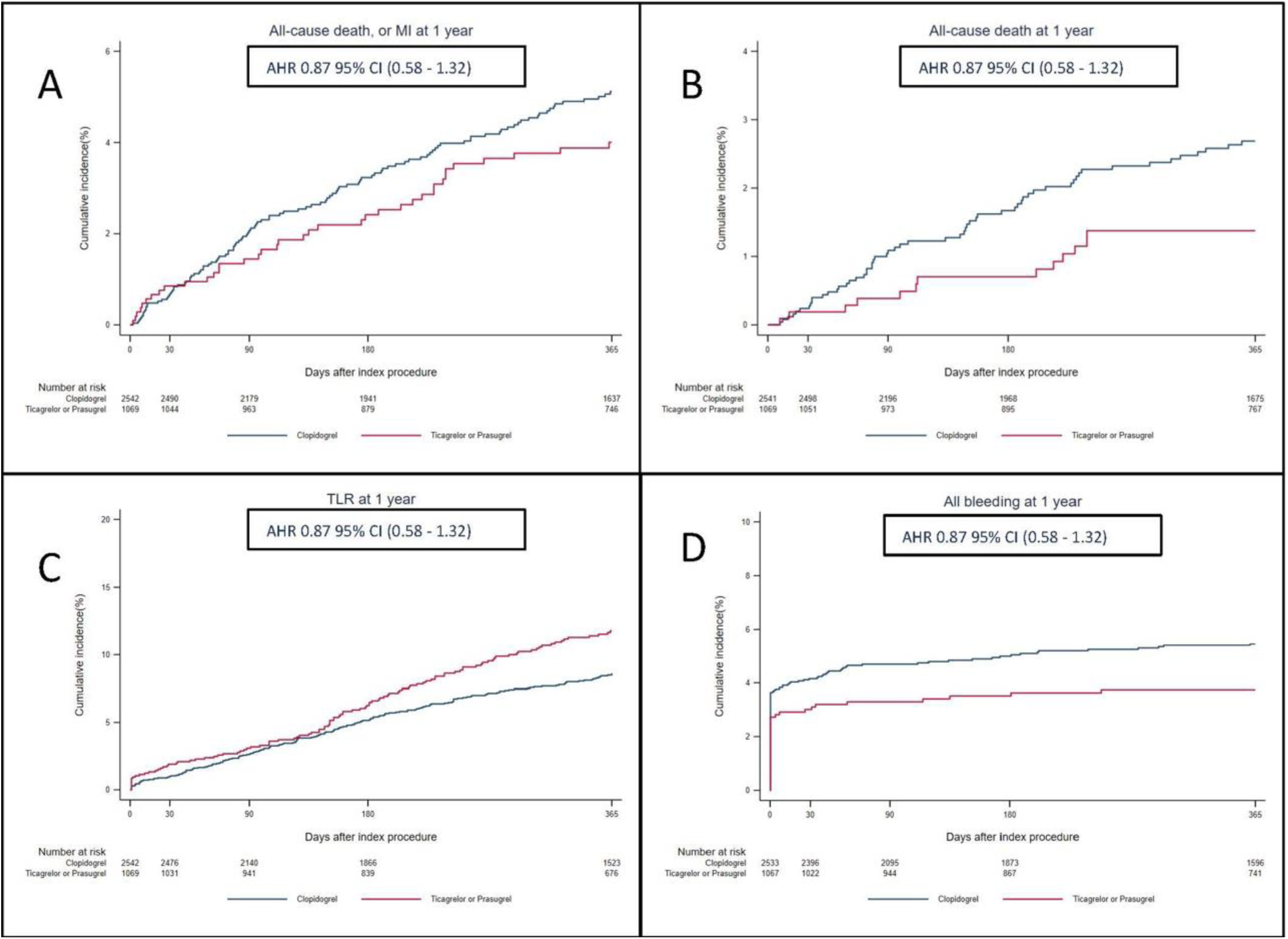
Clinical outcomes of ticagrelor or prasugrel with clopidogrel. (A) Composite outcome of death or myocardial infarction (MACE), (B) All cause mortality, (C) Target lesion revascularization (TLR) and (D) Bleeding at 1-year.

At 1-year follow up, patients treated with potent P2Y_12_ inhibitors were more likely to experience TLR on adjusted propensity score matching (16.0% vs 11.1%, AHR 1.26, 95%CI 1.01-1.57, p=0.045). A similar rate of major bleeding (3.7% vs 5.5%, AHR 0.98, 95%CI 0.66-1.46, p=0.923) and probable or definite stent thrombosis (0.4% vs 0.6%, AHR 0.54, 95%CI 0.17-1.71, p=0.296) was observed in both groups. The rate of CVA at 1-year follow up did not differ significantly between groups (0.5% vs 0.1%, AHR 3.00, 95%CI 0.66 - 13.6, p=0.155)

## DISCUSSION

Our real-world analysis comparing clopidogrel with potent P2Y_12_ inhibitors in patients undergoing PCI with atherectomy demonstrated several key findings. Firstly, our study demonstrated a significant increase in the use of potent P2Y_12_ agents over the study period with inherent differences in the patient populations who were prescribed either clopidogrel or ticagrelor/prasugrel. Second, risk-adjusted incidence of MACE at 1-year was not significantly different between patients prescribed a potent P2Y_12_ inhibitor compared to clopidogrel. Third, there was an equivalent risk of bleeding in both the potent P2Y_12_ and clopidogrel groups on adjusted analysis at 1-year follow-up. These findings provide real-world factors affecting clinicians’ choice of ticagrelor/prasugrel versus clopidogrel as well as their comparative efficacy and safety in a contemporary cohort of patients undergoing PCI with atherectomy.

Recent administrative data indicate that ticagrelor and prasugrel are used off-label in up to 30% of patients undergoing PCI for non-ACS indications.^11^ In our atherectomy patient cohort, it is notable that almost 40% of patients received ticagrelor or prasugrel between 2019-20 with a consistent increase in their use over the study period. The increasing tendency of clinicians to use P2Y_12_ inhibitors over time is likely multifactorial and may be due to supportive pharmacodynamic data, increasing physician familiarity with these medications as well as data extrapolated from the ACS cohorts.^3,11^ Moreover, implicit bias likely plays an important role in decision-making given the absence of supporting data for prescribing potent P2Y_12_ inhibitors in CCD thus far.

Consistent with data from the National Cardiovascular Data Registry (NCDR) Practice Innovation and Clinical Excellence (PINNACLE) registry,^3^ PCI complexity and a history of prior MI were all significant predictors for the use of ticagrelor/prasugrel use over clopidogrel, while advancing age was associated with clopidogrel use. Co-prescribing aspirin and ticagrelor in patients with prior MI (> 1 year) was previously examined and found to reduce the risk of MACE (MI, stroke, death) in the PEGASUS-TIMI 54 trial.^12^ However, this reduced risk of MACE came at the cost of significantly higher rates of Thrombolysis in Myocardial Infarction (TIMI) major bleeding,^4^ For ACS patients treated with potent P2Y_12_ inhibitors, the PLATO trial reported no increase in the overall risk of TIMI major bleeding but an increased risk of non-procedural bleeding with ticagrelor when compared to clopidogrel.^13^ Conversely, in the TRITON-TIMI-38 trial prasugrel was associated a significantly increased risk of TIMI major and life-treating bleeding.^5^ More recently, the ISAR-REACT 5 trial reported comparable bleeding risk in ACS patients treated with ticagrelor or prasugrel in conjunction with aspirin. ^14^ Importantly, unlike the TRITON-TIMI- 38 trial, prasugrel was only administered after the coronary anatomy was known, which is in keeping with our practice for CCD patients undergoing PCI. Reassuringly, the use of potent P2Y_12_ inhibitors was not associated with an increased risk of bleeding during follow-up in this study, although this is likely confounded by careful patient selection for potent P2Y_12_ inhibitors (younger, fewer patients with CKD and anemia, etc.). ^5^

We report a significantly higher rate of TLR in patients prescribed a P2Y_12_ inhibitor in conjunction with aspirin. This is likely reflective of unmeasured confounding variables in addition to procedure characteristics that were more prevalent in this group (bifurcation disease, CTOs, greater stent length, lower minimum lumen diameter) rather than the type of P2Y12 inhibitor co-prescribed with aspirin. Moreover, like previous studies, patient characteristics that predict TLR (diabetes mellitus, younger age, chronic kidney disease) were significantly more common in the potent P2Y_12_ inhibitor group of this study. ^15,16^ Furthermore, the observed similar rate of stent thrombosis at 1-year follow is in keeping with this increase in TLR not being a platelet-mediated complication. Finally, in contrast to the two pivotal trials demonstrating the benefit to using ticagrelor and prasugrel rather clopidogrel in the ACS population, there was no significant difference in the rate of MI during follow-up between groups in this study.^5,13^ Thus, the reduction in MI rates seen with using potent P2Y_12_ inhibitors in ACS was not reproducible in this CCD cohort and contributed to the overall similar rates in MACE between groups. The benefit of potent P2Y_12_ inhibitors in ACS and not CCD may be partially explained by significant endothelial disruption and inflammation in addition to sustained platelet hyperreactivity and thrombin generation after ACS. ^17^

Studies analyzing the use of potent P2Y_12_ inhibitors in CCD have focused mainly on periprocedural complications to date. In the randomized ALPHEUS trial, there was no reduction in periprocedural myocardial necrosis when ticagrelor was used in place of clopidogrel. ^18^ Furthermore, in the randomized SASSICAIA trial biomarker negative stable and unstable angina did not find a benefit to loading with prasugrel rather than clopidogrel in terms of 30-day MACE. ^19^ An observational study from China did report a significant reduction in MACE (cardiac death, MI, TLR) in patients undergoing PCI for complex stable CAD treated with ticagrelor rather than clopidogrel as part of DAPT. However, this study has inherent differences to our study due to being conducted in a Chinese population with only a small proportion of patients treated with potent P2Y_12_ inhibition (17.7%). Furthermore, MACE was driven by lower rates of cardiac death limiting direct comparison with our study.^20^

In the present study, we sought to evaluate whether the choice of P2Y12 agent would have an impact on cardiovascular events following coronary atherectomy. Our data indicated no significant differences in propensity adjusted MACE when comparing those patients who received clopidogrel or ticagrelor/prasugrel. These findings are in line with the existing data in our previously published all-comer CCD population and further support the use of clopidogrel over potent P2Y_12_ agents in patients undergoing more complex CCD requiring adjunctive atherectomy. ^21^ Thus, there remains a dearth of evidence for using potent P2Y_12_ inhibitors in CCD, and societal guidelines are unlikely to change without future studies (e.g., TAILORED-CHIP trial - tailored vs. conventional antiplatelet regimen for complex PCI) demonstrating clinical benefit and justification of the increased cost to patients when taking these antiplatelet agents. ^22^

### Limitations

Our study had notable limitations. Due to the observational design, it was difficult to establish causal relationships. Although a propensity score matched analysis was performed, there is a potential risk of bias secondary to unmeasured confounders not captured by the database affecting physician preference of prescription of potent antiplatelet versus clopidogrel. Furthermore, our database does not capture data on patient adherence after hospital discharge. Our institutional dose of prasugrel differs from conventional dosing as described above.^14^ However, the modified dose of prasugrel has shown to have similar efficacy with a lower risk of bleeding in certain patient populations ^23,24^. Lastly, we utilised all-cause mortality, not cardiovascular mortality in the assessment of the primary outcome, since the information on the cause of death is not available in this registry. By using all-cause mortality, we reduce the likelihood of introducing bias and adhere to the guidelines set forth by the Academic Research Consortium-2 consensus recommendations.

## CONCLUSION

In a cohort of patients with CCD undergoing PCI with atherectomy, ticagrelor/prasugrel had an overall similar impact on ischemic and bleeding complications compared to clopidogrel when co-prescribed with aspirin, except for a slight increase in TLR. Whether the use of potent antiplatelet agents has a role in mitigating the risk of adverse cardiovascular events in different subsets of the CCD population, such as those with a high thrombotic and low bleeding risk warrants further study.

## Data Availability

The data that support the findings of this study are available from the corresponding author upon reasonable request

## Non-standard Abbreviations and Acronyms

CCD: Chronic coronary disease
CSI: Cardiovascular solution inclusive
CTO: Chronic total occlusion
ELCA: Excimer laser coronary atherectomy
OA: Orbital atherectomy
RA: Rotational atherectomy
TLR: Target lesion revascularization

